# Additional therapies after minimally invasive treatments in women with OAB

**DOI:** 10.64898/2026.07.29.26359239

**Authors:** Yu Zheng, Mahir Maruf, Franco A Simonato, Whitney K Hendrickson, David Sheyn, James Hokanson, Caitlin Seibel, J Quentin Clemens, Aruna Sarma, Giulia M Ippolito

## Abstract

**Objective:** To evaluate the rate, timing, and predictors of additional therapy among women with idiopathic overactive bladder (OAB) following initial minimally invasive therapies (MIT).

**Study Design:** Retrospective single center cohort study of women with idiopathic OAB treated between 2012 and 2021. Using ICD and procedural codes, we identified women who underwent posterior tibial nerve stimulation (PTNS), sacral neuromodulation (SNM), or intradetrusor onabotulinumtoxinA (BTX). The primary outcome was receipt of additional OAB treatments including OAB medication and/or a different MIT after initial clinical response to MIT. Kaplan-Meier analysis estimated time to additional therapies; Cox proportional hazards and random survival forest models identified predictors.

**Results:** 1,007 women were included (PTNS: 459; SNM: 192; BTX: 356). At three years, 68% of PTNS patients, 52% of BTX patients, and 40% of SNM women required additional therapies with most patients receiving a prescription for additional pharmacotherapy rather than receiving a different MIT. Median time to additional treatments was 12 months (PTNS), 30 months (BTX), and 53 months (SNM). Multiple indications for SNM were associated with decreased hazard for AT (HR 0.56 95% CI 0.33-0.95, p=0.03) while recurrent urinary tract infections were associated with needing additional therapies in the BTX cohort (HR 1.8, 95% CI: 1.1-3.2).

**Conclusions:** Following initial MIT for idiopathic OAB, many women required additional treatment within three years, with most receiving a prescription for pharmacotherapy rather than receiving another MIT.

**Why This Matters:** Overactive bladder (OAB) affects over 33% of women in the United States, and current AUA/SUFU guidelines have shifted from traditional stepwise therapy toward shared decision-making and multimodal treatment. While minimally invasive therapies (MIT) are effective options for OAB, real-world data on patient trajectories after MIT initiation, including rates of treatment augmentation, crossover, or return to pharmacotherapy, remain limited.

This retrospective cohort of 1,007 women with idiopathic OAB provides novel data on post-MIT OAB therapy, with 40–68% of patients requiring additional therapy within three years. We noted significant differences in baseline characteristics and treatment tractories between the groups of patients stratified by first MIT and could not directly compare rates of AT between MITs. Factors associated with additional therapy included recurrent urinary tract infections after BTX, whereas multiple indications for SNM were associated with decreased use of AT. Most women who required additional treatment received pharmacotherapy rather than crossing over to a different MIT.

These findings reinforce that OAB management is a dynamic, ongoing process rather than a one-time intervention, and that additional therapy is common. Clinicians can use these data to set realistic expectations during counseling, framing additional therapy as a common and anticipated part of the treatment trajectory rather than a treatment failure.

## Introduction

Urinary urgency, frequency, nocturia with or without urgency urinary incontinence (UUI), the symptoms of Overactive bladder (OAB), affect more than 33% of women in America [1]. Management options range from conservative approaches like behavioral changes and pelvic floor physical therapy to minimally invasive interventions [2]. The 2024 SUFU/AUA guidelines for idiopathic OAB have shifted away from the traditional stepwise therapy model, advocating instead for a shared decision-making approach between patients and clinicians [2]. Despite various treatment options, nearly half of the patients discontinue OAB pharmacotherapy within six months, and only 3% to 15% proceed to minimally invasive therapies (MIT) like posterior tibial nerve stimulation (PTNS), sacral neuromodulation (SNM), or intradetrusor botulinum toxin (BTX) injections [3–5]. While multimodal care strategies are advocated for in current guidelines [2], there is little literature on trajectories of treatment following initial, successful MIT. Specifically, there is very little pragmatic, outcome-based literature evaluating rates of patients with OAB requiring additional therapy (AT) and the treatment options patients undergo after initial MIT [6–8]. Understanding the proportion of patients with OAB symptoms requiring AT is critical for patient-centered counseling. This is especially true considering prior work showing patients and clinicians express frustration at the discordance between expected and actual outcome [9]. This study aimed to evaluate the rate of, time to, and predictors of AT after MIT in women with OAB. We hypothesized that AT after MIT is common and that rates of, time to, and predictors of AT may vary among women with OAB based on both patient and clinical characteristics.

### Study Design

After Institutional Review Board approval (HUM00202983), a custom, retrospective, single institution database including patients with idiopathic OAB diagnosed between July 1, 2012 - July 1, 2021 was created using International Classification of Diseases (ICD) codes (Supplemental Table 1). The database included all patient encounters as well as patient demographics, medical history, and surgical history. Using this database, we used common procedural terminology (CPT) codes to identify a cohort of patients who underwent SNM, PTNS, or BTX injections at our institution. We performed a random 10% sample chart review to ensure >90% accuracy for identification of patients with OAB undergoing MIT for the same based on our computational model. Cohort Derivation is available as supplemental data (Supplemental Figure 1).

Exclusion criteria included: males, those younger than 18 years of age, prior diagnosis of neurogenic bladder, lack of follow-up after MIT, and MIT not associated with an OAB diagnosis..

The primary outcome of interest was the rate of receiving AT after the initial successful MIT. We defined AT as receipt of a prescription for an OAB medication or an encounter for a different MIT. Successful MIT is defined for each cohort in the sections below, but in brief this was 13 or more encounters for PTNS, 2 or more encounters for BTX or Stage II SNM. We did not consider the continuation of the same MIT as additional therapy.. We defined OAB medications as anticholinergics (oxybutynin, oxytrol, solifenacin, tolterodine, fesoterodine, trospium, darifenacin) or Beta-3 agonists (vibegron or mirabegron). PTNS was not queried after initial SNM or BTX because in our cohort very few patients progressed from SNM or BTX to PTNS. Implantable Tibial Nerve Stimulation was not offered during the study time period.

Secondary outcomes included the median time to, and predictors of AT following initial MIT. Baseline was defined as the date of either the first PTNS encounter, stage 1 implantation for SNM, or first BTX injection encounter. We could not reliably discriminate AT from multimodal therapy among those with SNM, based on available data, and therefore did not examine multimodal therapy within this study. We grouped patients based on their first successful MIT therapy.

### Statistical methods

Patient demographics were reported using median and inter-quartile range (IQR) for continuous variables and count and percent for categorical variables. Time to AT curves were developed using Kaplan-Meier methods with a 95% confidence interval to estimate event rates at 1 and 3 years and median time-to-AT. Curves were created separately for each initial MIT cohort. OAB medication prescription, SNM reprogramming, testing, or removal, or BTX injection (for the SNM cohort), SNM placement (for the BTX cohort), SNM testing or placement or BTX injection (for PTNS cohort) were defined as events. Among those without events, patients were censored at the latest date among the following events: urology visit, or urology questionnaire completion. Time to AT comparisons used log-rank tests.

Cox proportional hazards (PH) regression models adjusting for prespecified covariates evaluated time to AT, using complete-case analysis. Separate Cox PH models were created for the SNM, BTX, and PTNS cohorts. Six clinically selected covariates were included in the Cox PH models: BMI (≤25, 25-30, 30-35, >35 kg/m^2^), previous history of recurrent urinary tract infection (UTI), history of diabetes mellitus, estrogen use by age (<60 years old, ≥60 years old with no estrogen (topical or systemic) prescription, ≥60 years old with an estrogen prescription), multiple indications for SNM (SNM cohort only due to the broader indications including non-urological ones, such as fecal incontinence), Charlson comorbidity score (CCS), and total number of urology encounters prior to initial therapy. Statistical analyses were performed in SAS 9.4. P < 0.05 was considered significant.

Random survival forest (RSF) models identified predictors of AT after MIT. Models tested resulted in poor model performance and therefore results are not presented.

### Posterior Tibial Nerve Stimulation Technique and Cohort

Within our institution, patients undergo 12 weekly 30-minutes sessions of PTNS using the Urgent PC (Laborie) device per the manufacturer’s recommendations. Patients with a ≥50% improvement in OAB symptoms after 12 sessions are offered monthly PTNS maintenance. We performed a subgroup analysis to evaluate the difference in time to AT for women who underwent ≤12 weekly sessions versus ≥13. Women who received maintenance ( ≥13 PTNS) were classified “responders” to the initial PTNS therapy, as the responders typically complete the initial 12 weekly courses of the initial PTNS followed by monthly PTNS maintenance.

### Sacral Neuromodulation Technique and Cohort

Within our institution, patients initially undergo an SNM trial via an advanced test (a lead placement under anesthesia and a 14 day trial) or a peripheral nerve evaluation (an office lead placement with a 7 day trial). Patients with ≥ 50% improvement in symptoms, as verified by a bladder diary, are then offered to proceed to implant (stage 2) using a Medtronic InterStim^TM^ device. SNM responders were defined as patients who underwent a PNE or Stage 1 procedure (CPT 64561) followed by a Stage 2 procedure (CPT 64590), or who had a combined Stage 1 and 2 SNM (CPT 64561 and 64590). Patients who underwent PNE or Stage 1 (CPT 64561) without a subsequent Stage 2 were classified as non-responders (Supplemental Figure 1). The conversion to stage 2 among eligible patients was 91%. Due to the high conversion rate, and paucity of data for non-responders, we did not separately report the trajectory of AT among those who were exposed to stage 1 but did not undergo stage 2 (non-responders).

### Onabotulinum A Therapy and Cohort

Patients who received BTX for overactive bladder receive a first dose of 100 units of onabotulinum toxin A (BOTOX® Abbvie) reconstituted in 10 mL of preservative-free saline. The following doses could be increased to 200 units if clinically appropriate, but we did not have available data for dose at time of BTX injection in order to discriminate between 100 vs 200 units.. The injections were administered via 10, 1 ml injections throughout the bladder wall.

Therapy was predominantly performed in the clinic with flexible cystoscopy. Patients with ≥ 50% improvement in OAB symptoms were scheduled to undergo repeat injections. A subgroup analysis compared the difference in time to ATs for women who underwent only 1 injection (non-responders) versus repeated injections (2 or more; responders).

## Results

Using our inclusion and exclusion criteria, we grouped women in the following cohorts based on their first MIT: 459 underwent PTNS, 192 underwent SNM Stage II, and 356 underwent BTX. There were marked baseline differences across the three cohorts in terms of age, BMI, estrogen use by age, comorbidities and prior urologic evaluation/treatment (Table 1). The median follow-up was 15 months (IQR 5-36) for the PTNS group, 31 months (IQR 7-62) for the SNM group and 23 months (IQR 8-40) for the BTX group. The majority of AT consisted of pharmacotherapy and crossover rates from an first MIT to an additional MIT were low (4% for SNM; 7% BTX; 19% PTNS).

**Table 1.** Patient Demographics. Abbreviations: PTNS, posterior tibial nerve stimulation; SNM, sacral neuromodulation; BTX, intradetrusor botulinum toxin A; IQR, interquartile range; Rx, prescription; BMI, body mass index; POPQ, pelvic organ prolapse quantification, rUTI, recurrent urinary tract infections

| Variable | PTNS (N = 459) | SNM (N = 192) | BTX (N = 356) |
| --- | --- | --- | --- |
| **Age at initial therapy, median (IQR)** | 70 (62 - 78) | 59 (49 - 69) | 66 (56 - 75) |
| **Race, n (%)** |  |  |  |
| *Black or African American* | 29 (6.4%) | 15 (7.8%) | 26 (7.3%) |
| *White or Caucasian* | 395 (86%) | 167 (87%) | 308 (87%) |
| *Other* | 31 (6.8%) | 10 (5.2%) | 18 (5.1%) |
| *Unknown* | 2 (0.4%) | 0 | 4 (1.1%) |
| **Estrogen use by age, n (%)** |  |  |  |
| *<60 years old* | 104 (23%) | 101 (53%) | 113 (32%) |
| *>=60 years old and No Estrogen Rx* | 139 (30%) | 61 (32%) | 156 (44%) |
| *>=60 years old and Estrogen Rx* | 216 (47%) | 30 (16%) | 87 (24%) |
| **POPQ Performed, n (%)** | 128 (28%) | 21 (11%) | 70 (20%) |
| **Current alcohol user, n (%)** | 197 (43%) | 85 (44%) | 148 (42%) |
| *Missing* | 2 (0.44%) | 0 | 2 (0.56%) |
| **Current illicit drug user, n (%)** | 18 (3.9%) | 13 (6.7%) | 15 (4.2%) |
| *Missing* | 5 (1.1%) | 0 | 12 (3.4%) |
| **Current smoker, n (%)** | 17 (3.7%) | 23 (12%) | 34 (9.6%) |
| *Missing* | 0 | 0 | 1 (0.28%) |
| **Diabetes, n (%)** | 141 (31%) | 49 (26%) | 123 (35%) |
| **Multiple indications for procedure, n (%)** | N/A | 83 (43%) | N/A |
| **Positive urine culture within 30 days, n (%)** | 40 (8.7%) | 1 (0.52%) | 27 (7.6%) |
| **Negative culture, n (%)** | 23 (5.0%) | 1 (0.52%) | 34 (9.6%) |
| **No cultures within 30 days, n (%)** | 396 (86%) | 190 (99%) | 295 (83%) |
| **rUTI, n (%)** | 103 (22%) | 18 (9.4%) | 31 (8.7%) |
| **Prior medication use, n (%)** | 369 (80%) | 123 (64%) | 282 (79%) |
| **Prior urodynamics, n (%)** | 80 (17%) | 104 (54%) | 143 (40%) |
| **BMI, median (IQR)** | 29 (25 - 35) | 31 (27 - 36) | 32 (27 - 39) |
| **Charlson Comorbidity Index, median (IQR)** | 2 (0 - 8) | 1 (0 - 4) | 2 (0 - 6) |
| **Prior urology encounters, median (IQR)** | 1 (0 - 3) | 6 (4 - 10) | 2 (1 - 4) |

Percentages are based on non-missing data

### Posterior Tibial Nerve Stimulation Cohort

Among PTNS patients, 164 (36%) received monthly maintenance sessions and were considered responders. Among responders, the median time to any AT was 12 (95% CI 10-19) months (Figure 1A). The AT rate was 51% at 1 year and 68% at 3 years after initial PTNS (Table 2). After initial PTNS, 91 received new OAB medications, 18 underwent BTX, and 8 underwent SNM. The median time to a new prescription was 14 months (95% CI 11-25).

**Figure 1.**
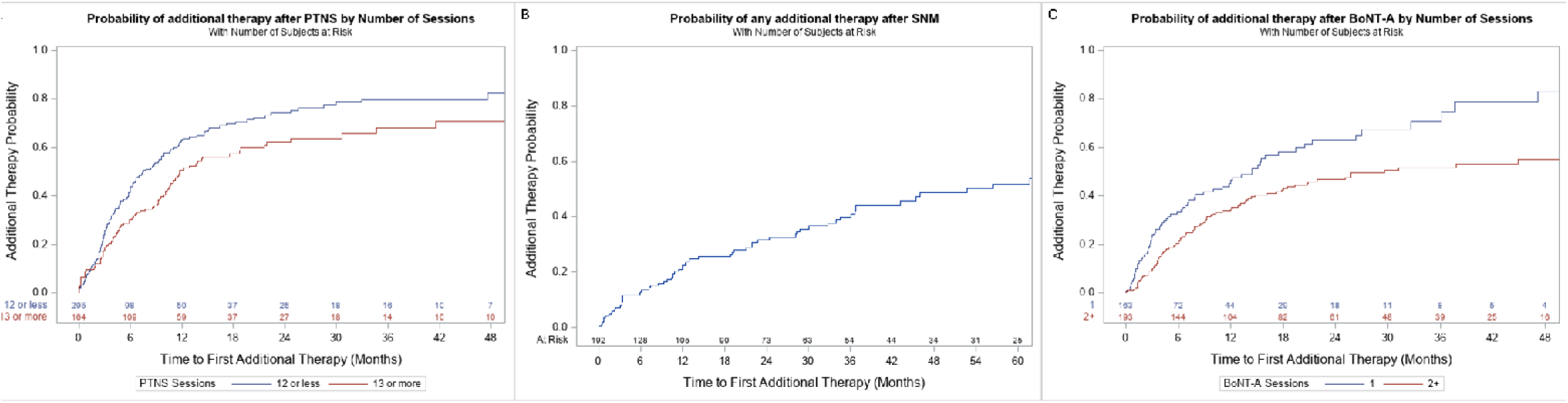
Kaplan-Meier curves for any additional therapy after index posterior tibial nerve stimulation (A), index Sacral Neuromodulation (B), or index intradetrusor botulinum toxin A (C)

**Table 2.** Rate of additional therapy, including additional medical therapy after the index procedure for overactive bladder, as calculated by Kaplan-Meier method.

| MIT |  | Rate of additional therapy (95% confidence interval) |  |  |  |  |  |  |  |
| --- | --- | --- | --- | --- | --- | --- | --- | --- | --- |
|  |  | 1-year follow-up |  |  |  | 3-year follow-up |  |  |  |
|  |  | Any additional therapy |  | Additional medical therapy |  | Any additional therapy |  | Additional medical therapy |  |
| **Posterior Tibial Nerve Stimulation**<br>(N at risk) | **Responders** | 51%<br>(59) | (43 - 59) | 47%<br>(64) | (40 - 56) | 68%<br>(14) | (58 - 77) | 65%<br>(18) | (56 - 75) |
|  | **Non-responders** | 63%<br>(50) | (56 - 70) | 48%<br>(70) | (41 - 55) | 80%<br>(16) | (72 - 86) | 68%<br>(25) | (60 - 76) |
| **Sacral Neuromodulation**<br>(N at risk) | **Responders\*** | 21%<br>(105) | (15 - 28) | 20%<br>(106) | (15 - 28) | 40%<br>(54) | (32 - 49) | 36%<br>(57) | (28 - 45) |
| **Intradetrusor Botulinum toxin**<br>(N at risk) | **Responders** | 35%<br>(104) | (28 - 42) | 32%<br>(106) | (26 - 40) | 52%<br>(39) | (44 - 60) | 47%<br>(42) | (40 - 55) |
|  | **Non-responders** | 46%<br>(44) | (38 - 56) | 41%<br>(47) | (32 - 50) | 71%<br>(8) | (59 - 82) | 62%<br>(10) | (51 - 74) |

Median follow-up in SNM cohort was 31 months, compared to BTX (23 months) and PTNS (15 months)
\*Non-reponder rates too low to model in Kaplan-Meier separately

Among responders, Cox PH model found no significant patient predictors of AT (Table 3).

**Table 3.** Cox proportional hazard ratios for the outcome of any additional therapy after index Posterior Tibial Nerve Stimulation, index Sacral Neuromodulation for overactive bladder, or index Intradetrusor Botulinum toxin for overactive bladder.

| Covariate | HR | 95% Confidence Interval |  | p-value |
| --- | --- | --- | --- | --- |
| Posterior Tibial Nerve Stimulation |  |  |  |  |
| BMI (ref= 25 - 30) | - | - | - | 0.083*** |
| <=25 | 1.19 | 0.83 | 1.70 | 0.3 |
| 30-35 | 1.34 | 0.92 | 1.95 | 0.12 |
| >35 | 1.56 | 1.11 | 2.2 | 0.011* |
| Recurrent UTI (Yes vs No) | 1.10 | 0.82 | 1.477 | 0.5 |
| Diabetes (Yes vs No) | 0.93 | 0.68 | 1.26 | 0.6 |
| Estrogen use by age (ref= <60 years old) | - | - | - | 0.4*** |
| >=60 and No Estrogen Rx | 1.26 | 0.90 | 1.76 | 0.18 |
| >=60 and Estrogen Rx | 1.10 | 0.76 | 1.59 | 0.6 |
| Charlson Comorbidity | 1.00 | 0.97 | 1.02 | 0.8 |
| Number of Urology Encounters | 1.02 | 0.97 | 1.07 | 0.4 |
| Sacral Neuromodulation |  |  |  |  |
| BMI (ref= 25 - 30) | - | - | - | 0.050*** |
| <=25 | 0.96 | 0.40 | 2.30 | 0.9 |
| 30-35 | 2.16 | 1.12 | 4.15 | 0.021* |
| >35 | 1.06 | 0.53 | 2.13 | 0.9 |
| Recurrent UTI (Yes vs No) | 0.48 | 0.18 | 1.31 | 0.15 |
| Diabetes (Yes vs No) | 0.98 | 0.55 | 1.76 | 0.9 |
| Estrogen use by age (ref= <60 years old) | - | - | - | 0.16*** |
| >=60 and No Estrogen Rx | 1.16 | 0.64 | 2.11 | 0.6 |
| >=60 and Estrogen Rx | 2.00 | 0.98 | 4.07 | 0.055 |
| Multiple Indications (Yes vs No)** | 0.56 | 0.33 | 0.95 | 0.032* |
| Charlson Comorbidity | 0.97 | 0.91 | 1.03 | 0.3 |
| Number of Urology Encounters | 1.04 | 0.99 | 1.09 | 0.12 |
| Intradetrusor Botulinum Toxin A |  |  |  |  |
| BMI (ref= 25 - 30) | - | - | - | 0.9*** |
| <=25 | 0.99 | 0.58 | 1.67 | 0.96 |
| 30-35 | 0.85 | 0.54 | 1.34 | 0.5 |
| >35 | 1.01 | 0.68 | 1.50 | 0.96 |
| **Recurrent UTI (Yes vs No)** | **1.89** | **1.14** | **3.16** | **0.015\*** |
| **Diabetes (Yes vs No)** | **0.96** | **0.69** | **1.35** | **0.8** |
| **Estrogen use by age**<br>**(ref= <60 years old)** | - | - | - | 0.075*** |
| >=60 and No Estrogen Rx | 1.21 | 0.85 | 1.74 | 0.3 |
| >=60 and Estrogen Rx | 0.76 | 0.49 | 1.16 | 0.2 |
| **Charlson Comorbidity** | **0.99** | **0.95** | **1.02** | **0.5** |
| **Number of Urology Encounters** | **1.05** | **1.00** | **1.11** | **0.051** |

\*statistically significant at $\alpha=0.05$ level
\*\*only available for SNM cohort.
\*\*\* global p-value

### Sacral Neuromodulation Cohort

In the SNM cohort, out of 210 eligible participants with stage 1, 192 underwent stage 2 (91%) and the median time to any AT was 53 months (95% CI: 36-75; Figure 1B) with AT rates of 21% after 1 year, and 40% after 3 years (Table 2). After initial SNM, 57 patients received pharmacotherapy prescriptions, and 7 patients underwent BTX treatment during the follow-up period. The median time to a prescription was 67 months (95% CI 43-75). Explant occurred in 35 (18%) patients.

Adjusting for covariates, participants who had multiple indications for SNM had half the risk of requiring ATs (HR 0.56 95% CI 0.33-0.95, p=0.03; Table 3). A BMI of 30-35 compared to 25-30 (HR 2.16, 95% CI: 1.12-4.15, p=0.021) was associated with an increased hazard of requiring AT, however this did not hold for BMI >35 and given the wide confidence interval the true effect may not be clinically significant. No significant associations were found between recurrent UTIs or estrogen use by age and the need for OAB AT in the SNM cohort.

### Onabotulinum toxin-A cohort

Among BTX patients, 193 (54%) received more than one session and were considered responders. Among responders, the median time to any AT was 30 (95% CI: 18-81) months (Figure 1C), with AT rates of 35% after 1 year and 52% after 3 years. After initial BTX treatment, 82 patients received a prescription for an OAB medication, and 15 patients transitioned to SNM. The median time to a new medication was 52 months (95% CI 21-81).

Adjusting for covariates, previous history of recurrent UTI was associated with an increased hazard of needing AT (HR 1.89, 95% CI: 1.14-3.16; Table 3). No other factors were significantly associated with AT for BTX.

## Discussion

This large, single-institution cohort revealed that AT after MIT for treatment of idiopathic OAB is common. At one year, 58% of patients treated with PTNS, 39% of BTX and 21% of SNM received a prescription for a bladder anticholinergic or beta-3-agonist or underwent another MIT (Table 2). The median time to AT was shortest after PTNS (12 months), followed by BTX (30 months), and finally SNM (53 months), however significant differences in baseline demographics for each group prohibit direct comparisons. This contemporary data provides pragmatic estimates of the need for AT following initial MIT and highlights the dynamic approach to OAB treatment even after initial successful MIT.

AT may represent either multimodal therapy or transition to a new therapy, our study could not discriminate between these two scenarios. The need for AT may be driven by either symptom progression leading to lack of efficacy of an ongoing MIT requiring multimodal therapy or patient and clinician decision to stop a MIT to progress to a new MIT or re-evaluate medications due to inefficacy, time commitment, or logistical barriers to therapy.

We found the highest rates of AT among those who initially received PTNS.The rate of PTNS response in our cohort matches several studies that reported poor long-term adherence to maintenance PTNS with continuation rates of 30-42.9% at 1 year [10,11]. Although a poor response to PTNS does not predict a poor response to SNM, very few patients in our cohort (7% at 3 years) proceeded to SNM following PTNS [12]. This may be due to diverging preferences of patients electing PTNS versus SNM, or clinician practice patterns [13].

Our study shows that at 1 year post initiation of BTX 35% of patients had either a prescription for an OAB medication or different MIT and those with previous history of recurrent UTI were more likely to need these ATs. While prior studies showed durable subjective improvement in symptoms and quality of life over 3 years with few discontinuing BTX [14]. There is little data on multimodal therapy with medications or additional MIT following BTX. Initiation of AT could be due to patient preference, adverse effects, loss of BTX efficacy due to progression of symptoms or potentially antibody formation [15], or multimodal efforts to bridge with medications to increase the duration and interval between BTX injections. The association between UTIs and the need for subsequent treatment post-BTX is particularly notable and may serve as a counseling point, given that UTIs are a known risk factor following BTX injections. When compared to SNM, BTX has a significantly higher risk of UTIs occurring within the 6 months of injection (35% vs 11%) as well as the need for intermittent catheterization [16]. These adverse events may lead patients who undergo BTX to seek alternative management strategies for their OAB.

We found that women who underwent SNM as first MIT had rates of AT that align with reported sustained improvements in patient-reported metrics even 5 years after the procedure [17]. We also found that those with multiple indications for SNM (ie bowel incontinence, urinary retention and/or urinary urgency incontinence) had half the hazard of AT compared to those with only urologic indications for SNM. While studies indicate that the vast majority of patients progressing to SNM discontinue their OAB medication and remain on SNM as their sole treatment [18], we could not identify studies that have examined resuming AT or multimodal therapy for OAB as an adjunct to MIT. We speculate that the low rate of AT could be due to several reasons: those undergoing SNM may have opportunities to modulate the device (reprogramming) to improve efficacy prior to starting AT, have a strong preference against resuming medications or BTX or have a relative contraindications to anticholinergic, beta-3 agonists or BTX (for example incomplete bladder emptying). The weak association between higher BMI and need for AT after SNM may be due to challenges with placement of the SNM lead with higher BMI [19] or due to worse OAB symptoms among those with higher BMI [13]. This could be considered when counseling patients about treatment options.

Our data revealed that only a small proportion of patients crossed over from one MIT to another. The AUA/SUFU OAB guidelines recommend trying an alternative MIT if the initial MIT fails. Like ours, prior studies show that only 4–9% of patients undergo another MIT after selecting an initial MIT other than pharmacotherapy [20,21]. This low crossover rate may reflect individual preference factors, since outcomes remain favorable when patients do proceed with an alternative MIT [20,21]. For example, prior research has shown a success rate of 43% for BTX after failed SNM and 66% for SNM after failed or intolerable BTX [8,22]. The reasons patients choose not to proceed with an alternative MIT after initial success and subsequent symptom recurrence are understudied but we hypothesize they could be multifactorial, encompassing treatment-specific adverse effects, logistical burden, insufficient patient education, insurance/financial barriers, and treatment fatigue.

This study has several limitations that must be considered when interpreting our findings. First, rates of AT between the three cohorts cannot be directly compared, as each cohort represents a clinically distinct subgroup. These baseline differences may offer avenues for further exploration of pragmatic distribution of MITs. Next, our study was single institutional data from the medical record and did not capture prescriptions or MITs provided outside of our institution. We considered AT a surrogate marker suggesting worsening of symptoms following MIT requiring either a new therapy or multimodal therapy. While this assumption is reasonable, given that patients would only seek AT if they were sufficiently bothered by persistent or worsening symptoms, it does not capture the nuances of individual patient experiences, their subjective satisfaction with MIT or AT, their severity of OAB at baseline, or progression of disease. We did not include patient-reported outcomes or objective measures of efficacy such as voiding diaries or pad tests as a measure of patient response to MIT or AT, nor comorbid pelvic floor disorders such as prolapse. The definition of AT was receiving a prescription for a OAB medication or undergoing a different MIT after initial successful MIT, even if they might be two different clinical scenarios having distinct clinical meanings and implications. Incorporating PROs in future studies would provide a more comprehensive understanding of treatment efficacy from the patient’s perspective. Furthermore, we did not evaluate rates of patients undergoing PTNS after SNM or BTX, possibly causing an asymmetry in the evaluation of crossover among MITs, this was due to very low rates of PTNS after SNM/BTX in our cohort [23,24]. We included several clinically important covariates, but could not include all relevant covariates due to sample size limitations, for example we did not include insurance data in our models. We reported data on AT for responders to initial MIT in order to avoid immortal-time bias affecting non-responding patients, therefore our data cannot be generalized to non-responders and these groups should not be directly compared. Finally, our cohort may not fully represent a generalized population as it is derived from a single, academic institution which may differ from those in other healthcare settings or populations.

Taken together, these findings have significant implications for clinical care and researchers. Patient counseling within a shared decision-making approach could present AT as a common occurrence, even following MIT. It further provides much-needed data on the rate of ATs after MIT and this underscores that more research is needed to understand these high rates and to explore best practices for OAB.

### Conclusions

Our findings demonstrate that management of OAB is an ongoing process after initial successful MIT, with up to 68% of women requiring AT within 3 years, and the majority using pharmacotherapy rather than additional MIT. This reframes how clinicians counsel patients: additional therapy after initial successful MIT is common. The high rates of AT underscore a critical evidence gap regarding AT and prospective studies aimed at understanding the reasons for AT are necessary because our data did not explore causation of AT use and whether or not this represented extension of prior therapy, escalation of therapy or failure of MIT.

## Data Availability

All data produced in the present study are available upon reasonable request to the authors

## Acknowledgements

Monica Van Til & Stephanie Daignault-Newton

## Ethics of approval statement

This study was approved by IRB at University of Michigan.

