## Supplemental Material for "Additional therapies after minimally invasive treatments in women with OAB"

**Supplemental Table 1:** ICD 9 and 10 codes for overactive bladder
**Supplemental Figure 1:** Patient selection flowchart. A, SNM cohort; B, BTX cohort; C, PTNS cohort.

**Supplemental Table 1:**
ICD 9 and 10 codes for overactive bladder

| CODE | DEFINITION |
|---|---|
| **ICD9** | |
| **788.3** | urinary incontinence |
| **788.30** | urinary incontinence, unspecified |
| **788.31** | urge incontinence |
| **788.33** | mixed incontinence in both males and females |
| **788.39** | other urinary incontinence |
| **788.43** | nocturia |
| **596.51** | hypertonicity of bladder |
| **788.41** | urinary frequency |
| **596.5** | overactive bladder, other functional disorders of bladder |
| **788.2** | sensation of incomplete emptying |
| **788.63** | Urgency of urination |
| **ICD10** | |
| **N39** | Other disorders of the urinary system |
| **N39.4** | Other specified urinary incontinence |
| **N39.8** | other specified disorders of urinary system |
| **N39.9** | disorder of urinary system, unspecified |
| **N32.9** | bladder disorder, unspecified |
| **R39.14** | feeling of incomplete bladder emptying |
| **R35.1** | nocturia |
| **R35.0** | frequency of micturition |
| **R32.81** | Overactive Bladder |
| **N39.41** | Urge Incontinence |

**Supplemental Figure 1:** Patient selection flowchart. A, SNM cohort; B, BTX cohort; C, PTNS cohort

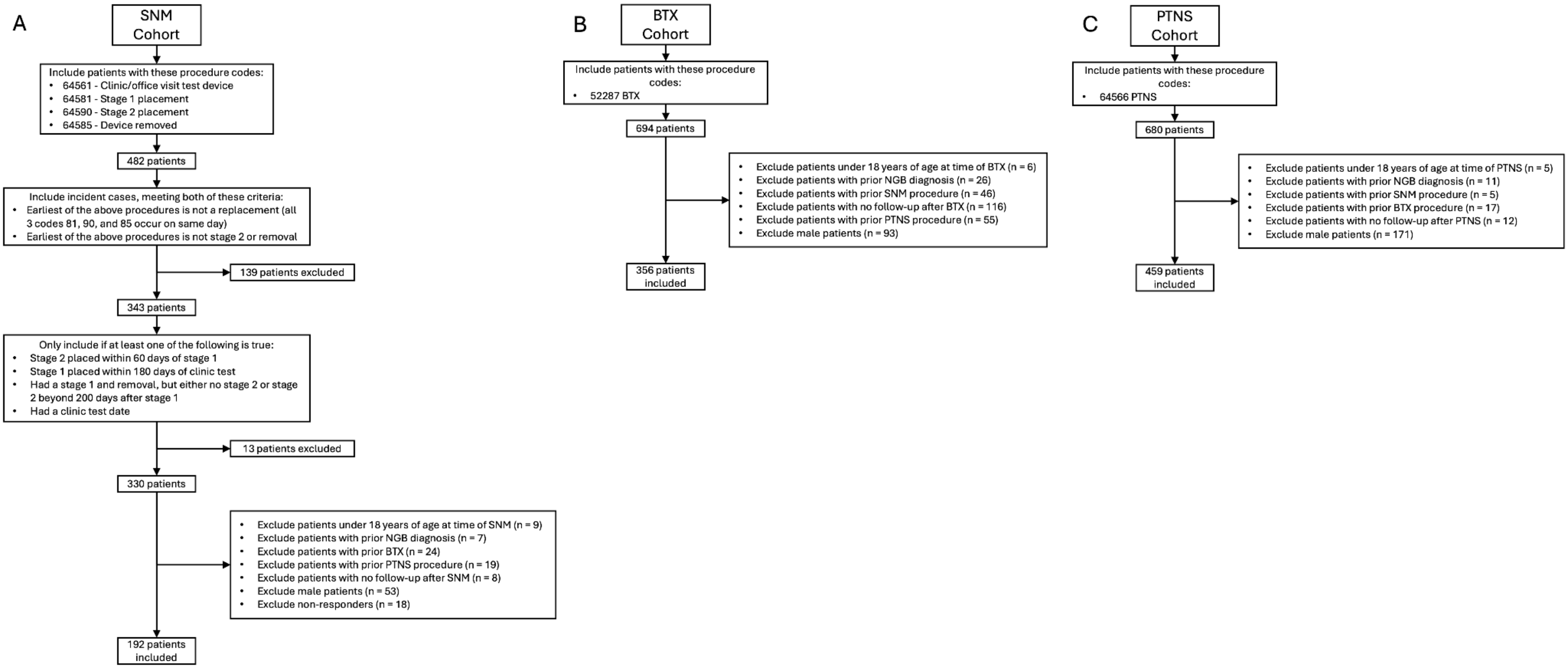
